# Evidence of long-lasting humoral and cellular immunity against SARS-CoV-2 even in elderly COVID-19 convalescents showing a mild to moderate disease progression

**DOI:** 10.1101/2021.02.23.21251891

**Authors:** Bastian Fischer, Christopher Lindenkamp, Christoph Lichtenberg, Ingvild Birschmann, Cornelius Knabbe, Doris Hendig

## Abstract

After the novel coronavirus severe acute respiratory syndrome coronavirus 2 (SARS-CoV-2) was first identified in China in late 2019, a pandemic evolved that has claimed millions of lives so far. While about 80 % of infections cause mild or moderate COVID-19 disease, some individuals show a severe progression or even die. Most countries are far from achieving herd-immunity, however, the first approved vaccines offer hope for containment of the virus. Although much is known about the virus, there is a lack of information on the immunity of convalescent individuals.

We here evaluate the humoral and cellular immune response against SARS-CoV-2 in 41 COVID-19 convalescents. As previous studies mostly included younger individuals, one advantage of our study is the comparatively high mean age of the convalescents included in the cohort considered (54 ± 8.4 years). While anti-SARS-CoV-2 antibodies were still detectable in 95 % of convalescents up to 8 months post infection, an antibody-decay over time was generally observed in most donors. Using a multiplex assay, our data additionally reveal that most convalescents exhibit a broad humoral immunity against different viral epitopes. We demonstrate by flow cytometry that convalescent donors show a significantly elevated number of natural killer cells when compared to healthy controls, while no differences were found concerning other leucocyte subpopulations. We detected a specific long-lasting cellular immune response in convalescents by stimulating immune cells with SARS-CoV-2-specific peptides, covering domains of the viral spike, membrane and nucleocapsid protein, and measuring interferon-*γ* (IFN-*γ*) release thereafter. We modified a commercially available ELISA assay for IFN-*γ* determination in whole-blood specimens of COVID-19 convalescents. One advantage of this assay is that it does not require special equipment and can, thus, be performed in any standard laboratory. In conclusion, our study adds knowledge regarding the persistence of immunity of convalescents suffering from mild to moderate COVID-19. Moreover, our study provides a set of simple methods to characterize and confirm experienced COVID-19.

## Introduction

The first cases of a novel respiratory disease occurred in Wuhan, China, in late December 2019. Polymerase chain reaction (PCR) testing identified the novel coronavirus severe acute respiratory syndrome coronavirus type 2 (SARS-CoV-2) early as a causative agent of the disease, which has been consequently named coronavirus disease 2019 (COVID-19) [1]. The virus has since spread worldwide and was classified as a pandemic by the WHO on March 11, 2020 [2]. While the first laboratory-confirmed case occurred in Germany on January 24, 2020 [3], the virus has so far infected millions of people worldwide. Two other coronaviruses with an increased pathogenic potential for humans have appeared in the last two decades: Severe acute respiratory syndrome coronavirus type 1 (SARS-CoV-1) occurred between November 2002 and June 2003 (8000 cases, 776 deaths) and Middle East respiratory syndrome coronavirus (MERS-CoV) in 2012 spread mainly on the Arabian Peninsula and infected 2428 people, 838 of whom died [4]. SARS-CoV-2 shows a sequence homology to SARS-CoV-1 of about 79.5 %, whereas only a homology of 50 % is found compared to MERS-CoV [5]. SARS-CoV-2 is an enveloped, single-stranded RNA virus, which is composed of four structural proteins: Spike (S), nucleocapsid (N), membrane (M) and envelope (E). Along with M and N, the E protein is responsible for the initiation and assembly of virus-like particles. Viral infection is realized by the binding of the trimeric S protein to the host cell’s angiotensin-converting enzyme 2 (ACE2) receptor. Thereby, the ectodomain of the S protein, which consists of the S1 subunit (containing the receptor binding domain: RBD) and the membrane fusion subunit (S2), plays a superior role [6]. The vast majority of infections are mild or even asymptomatic, however, the infection fatality rate is around 0.5 – 1 %, with the probability of fatal outcomes increasing with age [7]. The knowledge about a possible post-infection immunity is still limited and most studies include data from patients with more severe courses. It is assumed that both the humoral and cellular immune response have an impact on the severity. Most convalescents show detectable anti-SARS-CoV-2 IgG antibody levels between 10 and 21 days after infection [8]. However, there is evidence that some people, primarily those showing a mild progression, exhibit a delayed humorous response or even show no seroconversion at all [9]. According to recent studies, antibody persistence appears to depend primarily on the antibody class and COVID-19 severity. Anti-SARS-CoV-2 IgA and IgM antibody levels seem to drop rapidly, but IgG antibodies against the virus are detectable for several months in patients with a moderate or severe course [10, 11]. Furthermore, Wajnberg et al. found that more than 90 % of people who show general seroconversion also express neutralizing antibodies that can be detected for months and are directed primarily against the viral S protein [12]. Some publications suggest that the cellular immune response also plays an important role concerning SARS-CoV-2 containment and COVID-19 severity. Several studies show that a specific T-cell response in people with mild disease progression is triggered shortly after infection [13, 14]. By contrast, people with a severe course of the disease are more likely to show a dysregulated cellular immune response [15, 16]. Zuo et al. were able to demonstrate a robust T-cell response six months post-infection in all 100 initial seropositive individuals included in their study [17].

In our study, we characterized the cellular immune response of German convalescent blood donors suffering from mild to moderate COVID-19 compared to healthy blood donors without any history of SARS-CoV-2 infection. For this purpose, we modified a commercial enzyme-linked immunosorbent assay (ELISA) to rapidly and reliably measure the IFN-*γ* expression of leucocytes stimulated with a SARS-CoV-2-specific peptide pool covering different viral proteins. We examined the amounts of anti-SARS-CoV-2 IgA and IgG antibodies of convalescents over a period of up to eight months to assess the persistence of humoral immunity. We also provide detailed information on the profile of antibodies directed against different antigens of the new coronavirus using a novel multiplex bead-based assay.

## Methods

### Human Donors

The convalescents included in our study had a mild or moderate disease course not requiring hospitalization and SARS-CoV-2 RNA was initially detected by PCR. Blood samples of patients recovered from COVID-19 (*n* = 41; age: 54 ± 8.4; male: 57 %; female: 43 %) were obtained between 28 and 228 days after the onset of symptoms. An average of nine plasma donations were collected from each convalescent individual (range: 2 – 21 donations) during the study period. Healthy regular blood donors (*n* = 18; age 38 ± 13; male: 42 %; female; 58 %) with no known past or present COVID-19 disease served as controls. All probands of this study showed no signs of disease, such as fever or increased leukocyte counts, because potential blood donors with any suspicion of infection are excluded from donating blood. Characteristics of all donors are listed in Table S1.

All donors underwent a medical examination before donation. Samples were collected in accordance with the German Act on Medical Devices for the collection of human residual material. Ethical approval was obtained from the ethical committee of the HDZ NRW in Bad Oeynhausen (Reg.-No. 670/2020).

### Stimulation of immune cells using the SARS-CoV-2 peptide pool

An amount of 1 ml heparinized whole blood was treated with a SARS-CoV-2-specific peptide pool (Miltenyi Biotec, Bergisch-Gladbach, Germany) for immune cell stimulation. The peptide pool contained synthetic peptides whose sequences derived from the viral S, N and M proteins (final concentration of each peptide: 1 µg/ml). Treatment of whole blood with water served as a negative control. Stimulation with QuantiFERON Monitor (QFM) LyoSpheres (Qiagen, Hilden, Germany), which stimulate and activate different cell types of the innate and adaptive immune system, was used as a positive control. Samples were mixed by inverting and immediately incubated at 37 °C for 20 – 24 h. After incubation, samples were centrifuged (10 min; 2000 x g), plasma was collected and stored at -20 °C until use.

### Determination of interferon-*γ*in plasma

The QuantiFERON Monitor ELISA was used (Qiagen, Hilden, Germany) for the analysis of the IFN-*γ* amount in the plasma of controls and convalescents. The experimental procedure was conducted following the manufacturer’s instructions. The standard curve included different concentrations (8, 4, 2, 1, 0,25 and 0 IU/ml) of the IFN-*γ* standard supplied. Both peptide-stimulated and unstimulated samples (negative control) were measured undiluted. Standards and all donor-samples were measured in duplicate. All samples were subsequently diluted 1:1 with murine anti-human IFN-*γ* horseradish peroxidase (HRP), transferred to a 96-well plate coated with anti-human IFN-*γ* monoclonal antibodies and incubated for 2 h at room temperature (RT). After a washing step, enzyme substrate solution was added to each well. The plate was incubated for 30 min at RT followed by the addition of a stop solution. The measurement was performed at an optical density of 450 nm (reference at 650 nm) using a Tecan Reader Infinite-pro 200.

### Determination of anti-SARS-CoV-2 IgG and IgA antibodies (Euroimmun)

Two commercial ELISAs (Euroimmun, Lübeck, Germany) were used for the determination of anti-SARS-CoV-2 antibodies (IgG and IgA). The assays were conducted automatedly, following the provider’s instructions. Briefly, diluted plasma samples (1:101) and undiluted positive control, negative control and calibrator were incubated for 1 h at 37 °C in ELISA plates coated with recombinant-generated S1 domain of the viral S protein. After incubation and three consecutive washing steps, the plates were incubated with enzyme conjugate for 30 min at 37 °C. After another three washing steps, the plates were incubated with a substrate solution for 15 min at RT, followed by the addition of a stop solution. The optical density was measured at 450 nm (reference at 620) using the Euroimmun Analyzer I system.

### Measurement of anti-SARS-CoV-2 S1/S2 IgG antibodies (DiaSorin)

The LIAISON SARS-CoV-2 IgG chemiluminescent assay is designed to measure IgG antibodies against the S1 and S2 subunits of the viral S protein. The automated assay format consists of an initial incubation (10 min) of S1/S2-coated paramagnetic microparticles (PMPs) with 20 µl of test-serum diluted in assay buffer. After a washing step, polyclonal ABEI-labeled goat anti-human IgG is added to the PMPs, the mixture is incubated for another 8 min and washed again. Finally, starter reagents are added and the relative light units (RLU) emitted, which are proportional to the anti-SARS-CoV-2 IgG levels, are converted to arbitrary units (AU/mL) according to a standard curve.

### Multiplex assay for the differentiated detection of anti-SARS-CoV-2 antibodies

The LABScreen COVID Plus multiplex assay (ThermoFisher, Waltham, USA) was used to determine the expression pattern of anti-SARS-CoV-2 antibodies directed against differential viral epitopes (within the S, S1, S2, RBD and N protein). The SARS-CoV-2 antigens are immobilized on microbeads, each of which contains a unique dye signature which was separately detected using a Luminex 200 flow analyzer. The assay was performed according to the manufacturer’s instructions. Briefly, control and test serum were diluted 1:10 in 1 x phosphate-buffered saline (PBS) /0.02 M ethylenediaminetetraacetic acid (EDTA). An amount of 20 µl of the diluted serum was mixed with 5 µl of the LABScreen beads, each entire volume was transferred into a separate well of a 96-well plate and incubated for 30 min in the dark with gentle shaking. After the samples had been washed three times with 200 µL of a 1 x wash solution, the plate was centrifuged at 1300 g for 5 min and the supernatant was discarded. An amount of 100 µl of 1X phycoerythrin-conjugated anti-human IgG was added to each well and the plate was again incubated for 30 min in the dark with gentle shaking. After the samples had been washed twice more, 80 µl of 1 x PBS was added to each well of the plate. Data was acquired and analyzed using the following formula:

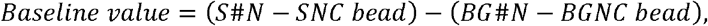

using the following definitions:

**S#N** Sample-specific fluorescent value for bead #N

**SNC bead** Sample-specific fluorescent value for negative control (NC) bead

**BG#N** Background NC serum fluorescent value for bead #N

**BGNC bead** Background NC serum fluorescent value for NC bead

**NC Serum** Negative control serum

### Determination of neutralizing antibodies against SARS-CoV-2

The determination of neutralizing antibodies against SARS-CoV-2 was applied by using the cPass™ SARS-CoV-2 Neutralization Antibody Detection KIT (GenScript, Piscataway Township, USA). Experimental procedure was performed following the manufacturer’s instructions. Briefly, EDTA coagulated blood was centrifuged (10 min, 2000 x g), EDTA plasma was collected and measured directly or stored at -20 °C. Controls (positive, negative) and plasma samples were measured in a 1:10 dilution. All samples and controls were mixed with a *horseradish peroxidase*-RBD solution and incubated at 37 °C for 30 min. Afterwards, samples and controls were incubated on plates coated with ACE2-receptor for 15 min at 37 °C. After incubation and washing steps, the substrate solution was added to each well and the plate was incubated for 15 min at RT followed by the addition of the stop solution. Optical density was measured immediately at 450 nm with the Tecan Reader Infinite-pro 200. The inhibition capability was calculated as follows:

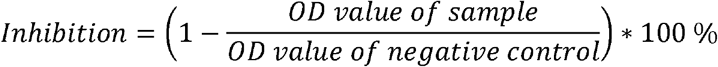

According to the manufacturer, values > 20 % were considered positive concerning neutralizing antibodies.

### Flow Cytometry

The EDTA blood was analyzed by flow cytometry to determine the lymphocyte subpopulations and immune status of donor blood. Consequently, samples were incubated with a mixture of fluorochrome-tagged monoclonal antibodies (BD Multitest™ 6-Color TBNK; Table 1) for 15 min at RT in the dark. Afterwards, each sample was mixed with 500 µl FACS Lysing solution (BD Biosciences, Franklin Lakes, USA) to remove erythrocytes and again incubated for another 15 min at RT in the dark. After incubation, samples were analyzed using a BD FACS Canto™ II device. A total of 3000 individual cells were recorded for each sample using a dot plot combination of low angle forward scattered (FCS) and right angle scattered (SSC) laser light. Data were analyzed using BD FACS DIVA software.

**Table 1:**
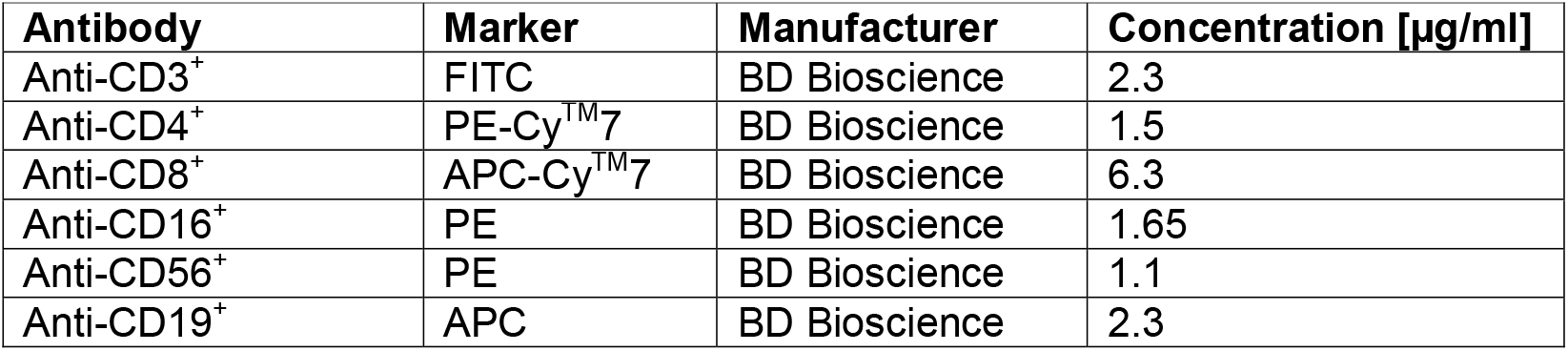
Antibodies used for the analysis of lymphocyte subpopulations.

| **Antibody** | **Marker** | **Manufacturer** | **Concentration [µg/ml]** |
| --- | --- | --- | --- |
| Anti-CD3<sup>+</sup> | FITC | BD Bioscience | 2.3 |
| Anti-CD4<sup>+</sup> | PE-Cy™7 | BD Bioscience | 1.5 |
| Anti-CD8<sup>+</sup> | APC-Cy™7 | BD Bioscience | 6.3 |
| Anti-CD16<sup>+</sup> | PE | BD Bioscience | 1.65 |
| Anti-CD56<sup>+</sup> | PE | BD Bioscience | 1.1 |
| Anti-CD19<sup>+</sup> | APC | BD Bioscience | 2.3 |

The Sysmex XN-1000 (Sysmex, Kōbe, Japan) flow cytometer was used to determine white blood cells, lymphocytes and monocytes in donors’ EDTA blood.

## Results

### Increased IFN-*γ* release in COVID-19 convalescents after peptide stimulation

Figure 1 shows the IFN-*γ* release of the unstimulated and stimulated whole blood of convalescent donors and healthy controls.Treatment with specific SARS-CoV-2 peptides led to a highly significant increased IFN-*γ*-release (3.93 ± 0.71 IU/ml vs. 0.34 ± 0.10 IU/ml; p < 0.0001) in convalescents compared to unstimulated whole blood. The release of IFN-*γ* did not differ significantly between the stimulated and unstimulated whole blood of healthy controls. There was no significant difference in the IFN-*γ* secretion between the unstimulated whole blood of convalescents and controls. The IFN-*γ* release in the whole blood of stimulated convalescents was significantly increased compared to the stimulated plasma of control donors (3.39 ± 0.71 vs. 0.43 ± 0.14; p < 0.0001).

**Figure 1:**
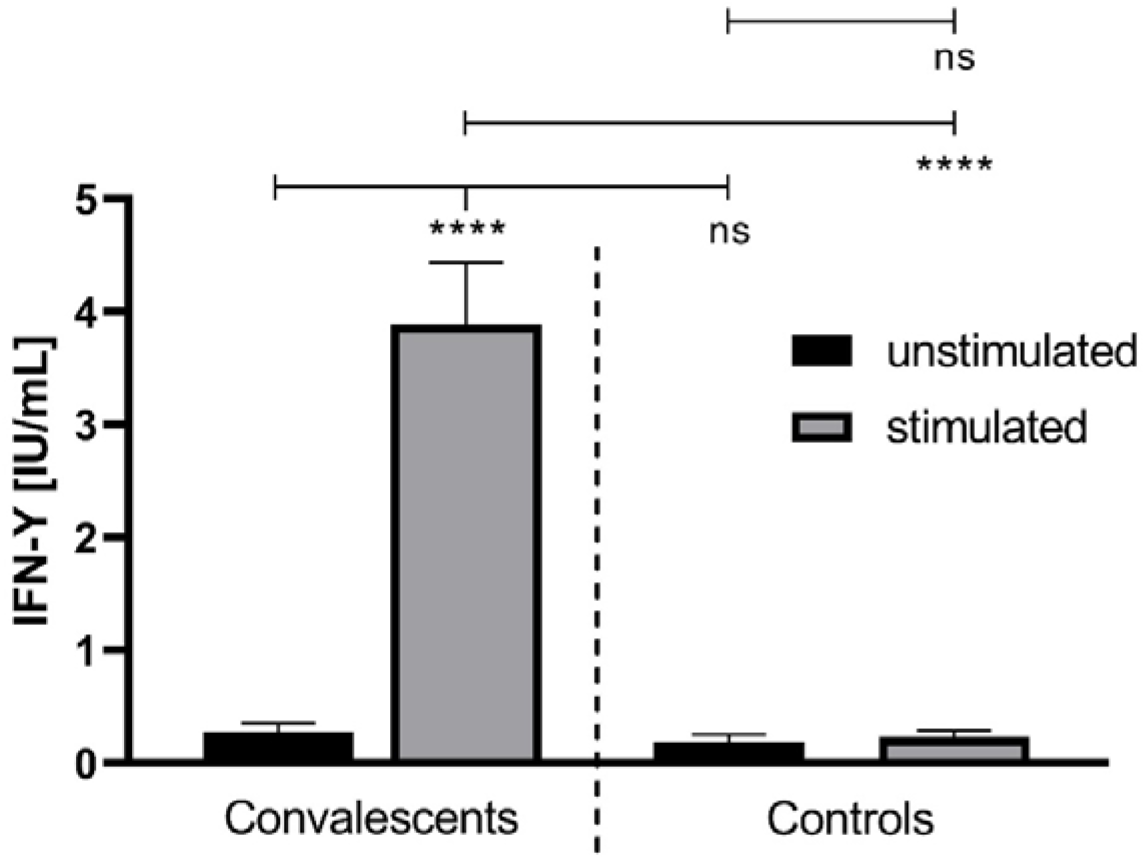
The IFN-*γ* concentration in unstimulated and stimulated whole blood of convalescent COVID-19 donors (n = 41) and healthy controls (n = 18). **The** IFN-*γ* release was monitored after treatment of the whole blood, donated from COVID-19 convalescents and healthy controls, with a SARS-CoV-2-specific peptide pool (grey bars). The latter contained synthetic peptides whose sequences derived from the viral spike (S), nucleocapsid (N) and membrane (M) proteins (final concentration of each peptide: 1 µg/ml). The treatment of whole blood with water served as a negative control (black bars). ****: p < 0.0001; ns: not significant (Mann Whitney U test).

### No correlation between IFN-*γ* concentration and anti-SARS-CoV-2 IgG expression

The IFN-*γ* release was correlated to the anti-SARS-CoV-2 IgG expression determined by both a semiquantitative ELISA assay (Euroimmun) and a quantitative CLIA test (DiaSorin). There was no significant difference in the IFN-*γ* release between the stimulated whole blood of convalescent donors showing low (IgG ratio 1.0 – 1.9), medium (IgG ratio 2.0 – 3.9) or high (IgG ratio > 4) anti-SARS-CoV-2 IgG antibody ratios using the Euroimmun assay, as seen in Figure 2A. Additionally, no significant differences in the IFN-*γ* release were detected between grouped (low: 0 – 30 AU/ml; medium: 31 – 100 AU/ml; high: > 100 AU/ml) anti-SARS-CoV-2 IgG concentrations in the quantitative DiaSorin assay (Fig. 2C). These observations were confirmed by linear regression, which showed only a weak correlation between the IFN-*γ* concentration and anti-SARS-CoV-2 IgG expressions determined using the Euroimmun (r = 0.2831, Fig. 2B) and DiaSorin (r = 0.2578, Fig. 2D) assay, respectively.

**Figure 2:**
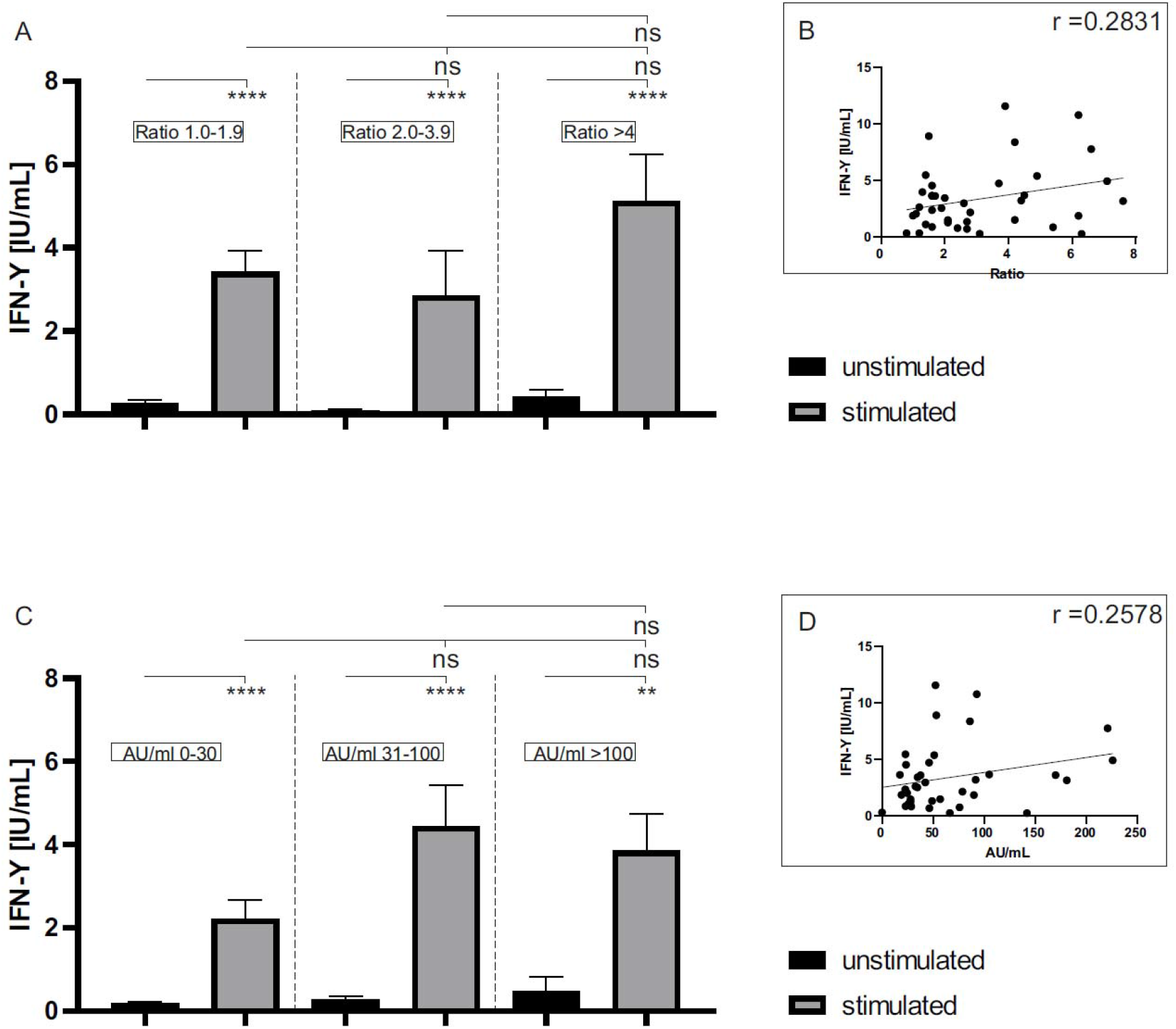
The IFN-*γ* release in unstimulated and stimulated whole blood of COVID-19 convalescent donors (n = 41) in relation to anti-SARS-CoV-2 IgG levels determined using two different serological assays. Anti-SARS-CoV-2 antibodies of convalescent donors, determined by the semiquantitative Euroimmun (A) or the quantitative DiaSorin (C) assay, were grouped and related to the IFN-*γ* release of unstimulated (black bars) and stimulated (grey bars) whole blood. A pool of synthetic peptides whose sequences derived from the viral S, N and M proteins (final concentration of each peptide: 1 µg/ml) was used for stimulation. Water treatment served as a negative control (unstimulated). In addition, the individual results of the antibody measurements were plotted against the respective IFN-*γ* release to perform a linear regression (B and C). ****: p < 0.0001; **: p < 0.002; ns: not significant (Mann Whitney U test).

### Multiplex assay for the qualitative detection of anti-SARS-CoV-2 IgG antibodies

The COVID-Plus multiplex assay was conducted to detect antibodies qualitatively against the S, S1, RBD, S2 and N of SARS-CoV-2. Antibodies against the viral S, S1, RBD and S2 protein were detected in all but one convalescent donors, as can be seen in Figure 3. Only 61 % (25/41) of those who recovered from COVID-19 expressed antibodies against the viral N. While all controls were generally negative for the expression of SARS-CoV-2 specific antibodies as expected, antibodies to the viral S protein were detected in one control donor.

**Figure 3:**
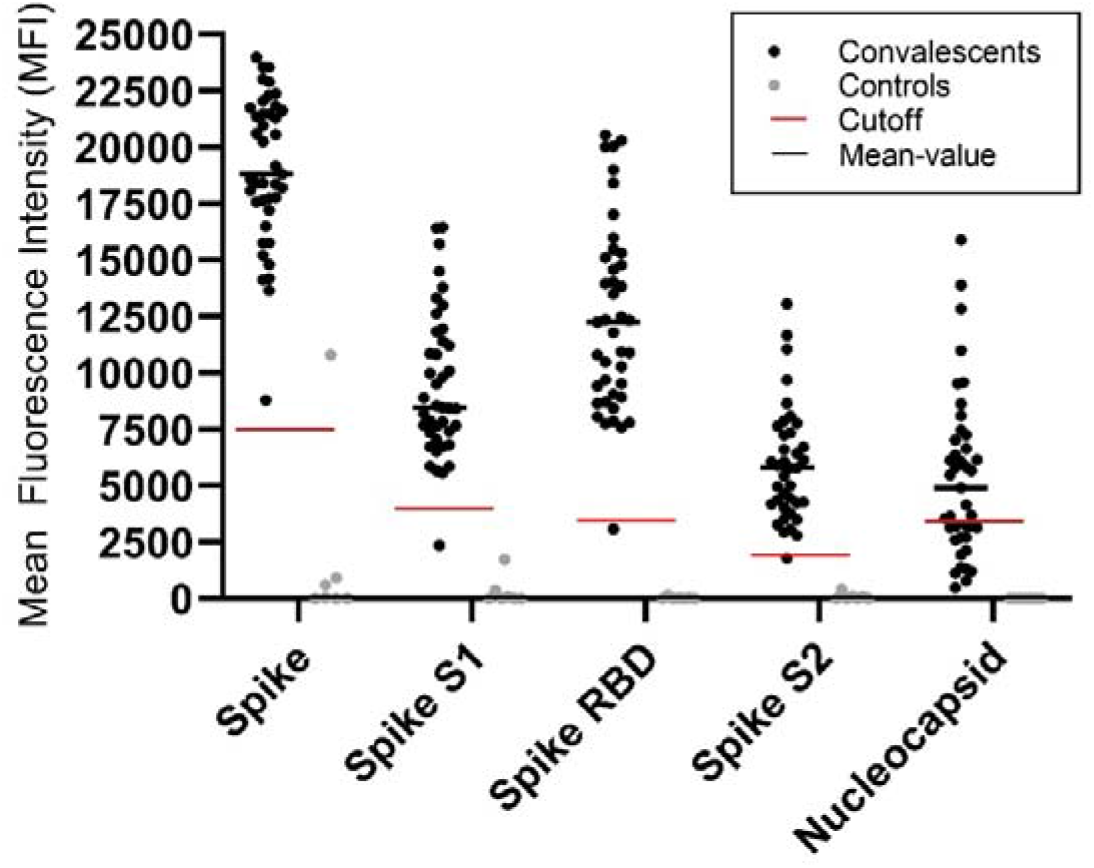
Qualitative detection of anti-SARS-CoV-2 antibodies in convalescents (n = 41) and healthy controls (n = 7) using a multiplex flow cytometry assay. The serum of convalescents and controls was used to identify antibodies binding to beads coated with various purified SARS-CoV-2 antigens. Samples were considered positive if they had a mean fluorescence intensity (MFI) value above the manufacturer’s cutoff. The individual cutoff for each antibody was set as follows: S: 7500; S1: 4000; RBD: 3500; S2: 1900; N: 3500.

### Expression of neutralizing anti-SARS-CoV-2 antibodies

The expression of neutralizing antibodies against SARS-CoV-2 was increased significantly in the plasma of convalescent donors (60.82 ± 3.15 % inhibition capability), as shown in Figure 4A. As might be expected, the inhibition capability in healthy controls was below the manufacturer’s cutoff of 20 % (12.23 ± 1.02 %).

**Figure 4:**
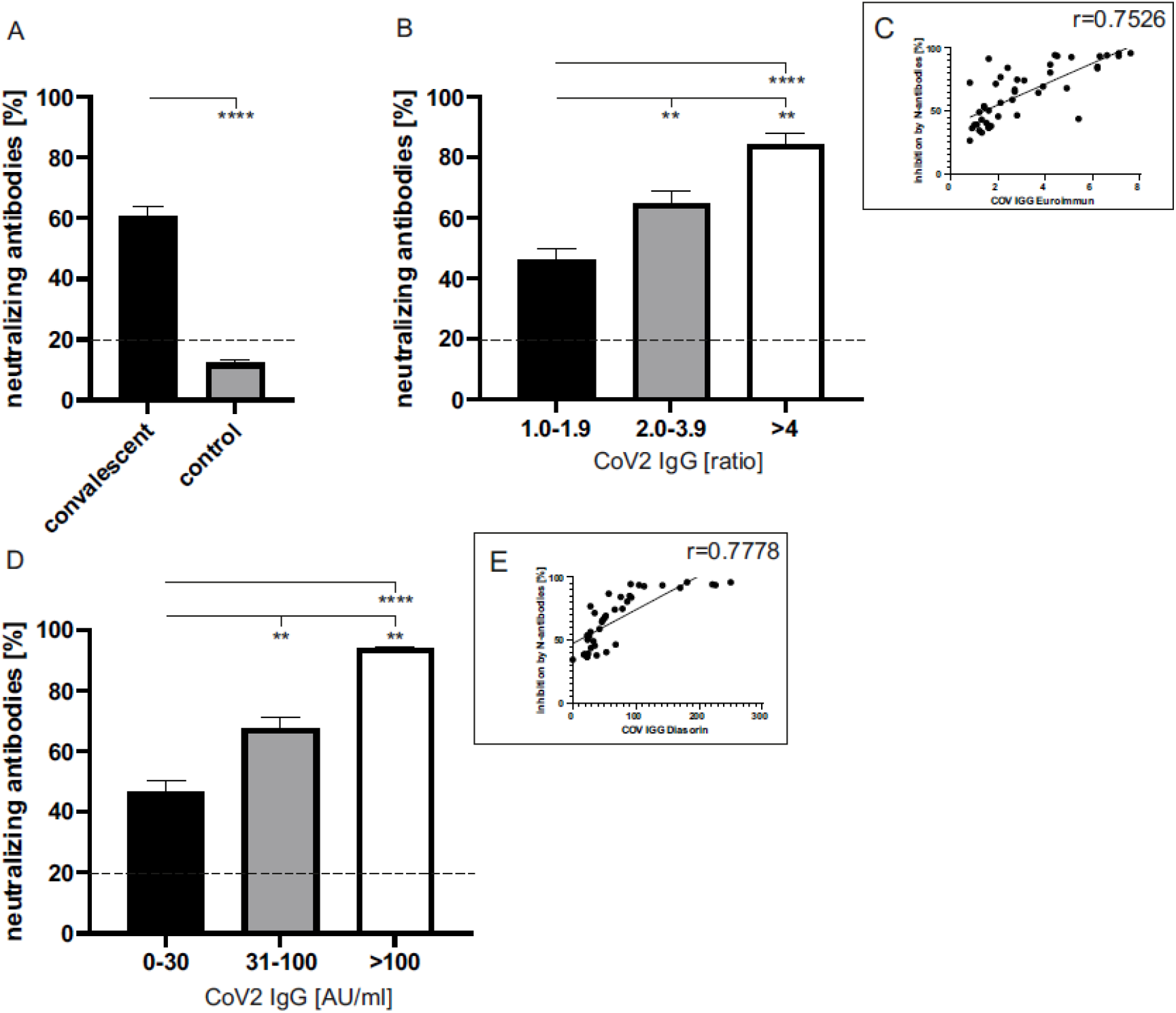
The inhibition capability of neutralizing antibodies in the plasma of convalescent COVID-19 donors (n = 41) and controls (n = 18). **(A)** The general inhibition capability of neutralizing antibodies in the plasma of convalescents (black bar) and healthy controls (grey bar). According to the manufacturer, the cutoff was set to 20 %. **(B)** Correlation between the inhibition capability of neutralizing antibodies and anti-SARS-CoV-2 IgG ratios (Euroimmun) in the plasma from convalescent donors. Expressions were grouped as follows: low (IgG ratio: 1.0 – 1.9), medium (IgG ratio: 2.0 – 3.9) and high (IgG ratio: > 4) **(B, C)**. Correlation between the inhibition capability of neutralizing antibodies and anti-SARS-CoV-2 IgG concentrations (DiaSorin) in the plasma from convalescent donors. Expressions were grouped as follows: low (0 – 30 AU/ml), medium (31 -1 00 AU/ml) and high (> 100 AU/ml) **(D, E)**. The dashed line symbolizes the manufacturer’s cutoff (< 20 %: negative; > 20 % positive). ****: p < 0.0001; **: p < 0.002 (Mann Whitney U test).

In addition, a moderate correlation between the neutralizing and general anti-SARS-CoV-2 IgG antibody expression, determined by using the Euroimmun (Fig. 4B and C, r = 0.7526) and DiaSorin (Fig. 4D and E, r = 0.778) assay, was detected. In detail, a significant increase of inhibiting neutralizing antibodies was shown in the plasma of convalescent donors with medium (Euroimmun: ratio 2.0 – 3.9; DiaSorin: 31 – 100 AU/ml) in comparison to those with low (Euroimmun: ratio 1.0 – 1.9; DiaSorin: 0 – 30 AU/ml) IgG antibody levels (65 ± 3.96 % and 67.42 ± 3.70 % vs. 46.48 ± 3.65 % and 46.55 ± 3.54 %, respectively). Convalescents showing high (Euroimmun: ratio > 4; DiaSorin: > 100 AU/ml) anti-SARS-CoV-2 IgG values in both assays tested also showed the highest inhibition capabilities (84.36 ± 3.51 % and 93.91 ± 0.53 %, respectively).

### Higher amount of natural killer cells in blood of convalescent donors

The composition of leucocytes in whole blood specimens of convalescents and healthy controls was analyzed by flow cytometry. There was no significant difference in the amount of immunological cell subpopulations between both groups, as shown in Figure 4, except of natural killer (NK) cells (Fig. 5D). When compared to controls (7.68 ± 1.03), the NK cell expression was increased significantly in COVID-19 convalescents (10.76 ± 0.80 %).

**Figure 5:**
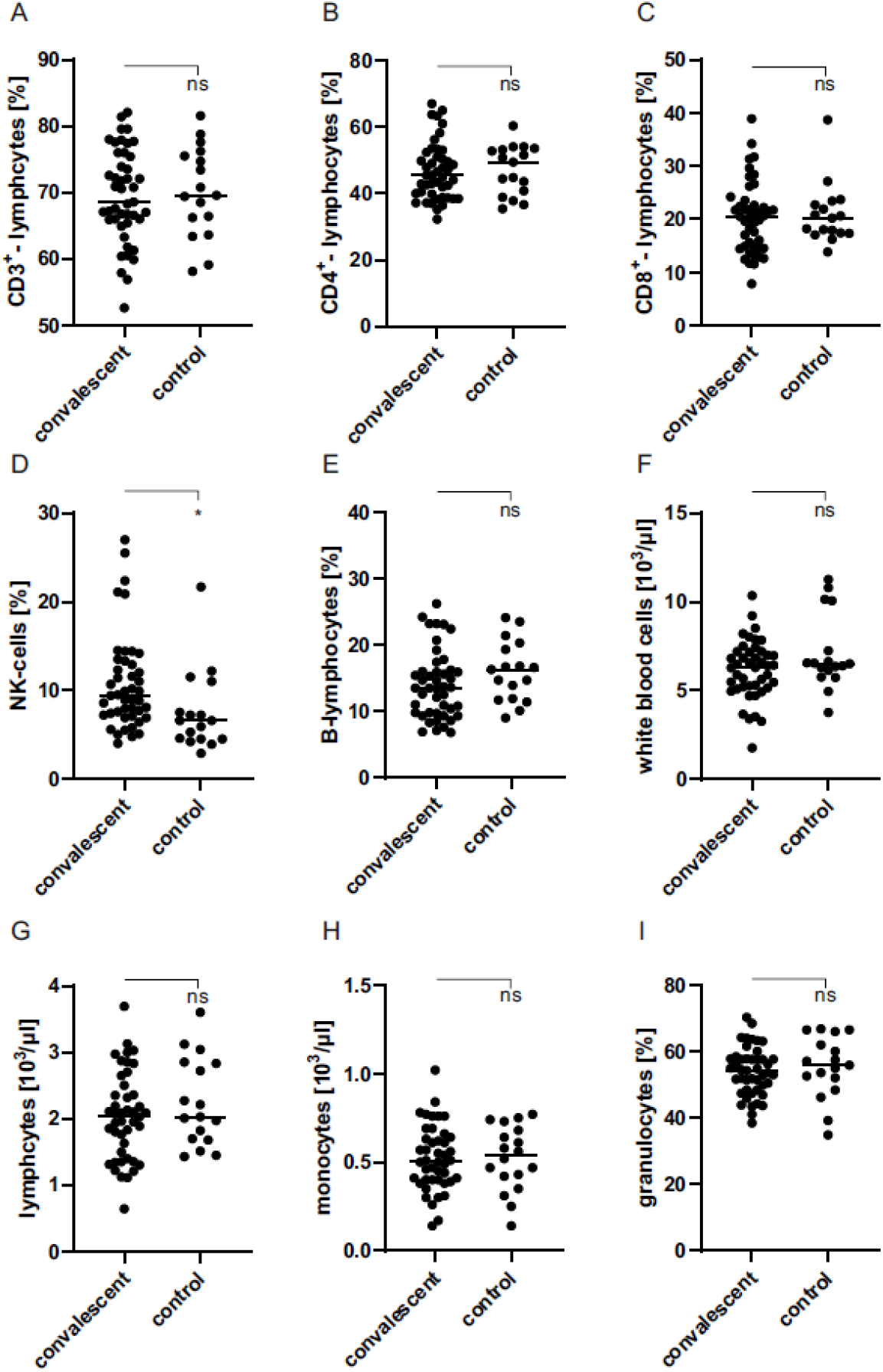
Determination of immunological cell subpopulations in whole blood specimens of COVID-19 convalescents (n = 41) and healthy controls (n = 18) by flow cytometry. Subpopulations: CD3^+^ lymphocytes **(A)**, CD4^+^ lymphocytes **(B)**, CD8^+^ lymphocytes **(C)**, NK cells **(D)**, B lymphocytes **(E)**, white blood cells **(F)**, lymphocytes **(G)**, monocytes **(H)**, granulocytes **(I)**. **: p < 0.002; ns: not significant (Mann Whitney U test).

## Discussion

The COVID-19 pandemic has claimed millions of lives so far worldwide. First vaccines have been developed and approved in record time,however, many questions regarding the immunity of convalescents to the SARS-CoV-2 virus remain unresolved. In this regard, most studies focus on the humoral immune response, although cellular immunity also appears to have an impact on the severity of the disease. We stimulated whole blood with a SARS-CoV-2-specific peptide pool (including sequences of the viral S, M and N protein) and subsequently determined IFN-*γ* release to evaluate the cellular immune response of convalescent and healthy control donors. To the best of our knowledge, we are the first to use a modified version of the QuantiFERON Monitor assay (Qiagen, Hilden, Germany) for this purpose. The assay is particularly suitable if, as in our study, a general statement about IFN-*γ* release before and after stimulation is made. In order to be able to make cell-specific assessments regarding the release of IFN-*γ*, exemplarily commercial ELISPOT systems or special flow cytometry-based assays should be used. Nevertheless, the ELISA-based assay used in our study shows important advantages concerning its rapid and simple feasibility, its commercial ready-to-use availability (no manual plate-coating required) and the fact that a standard microplate-reader and no special software are required for the analysis. This allows the assay to be performed by any laboratory with standard equipment. Cells in our study were stimulated in a parallel setup using LyoSpheres pellets containing a CD3 T-cell receptor agonist and a viral TLR 7/8 ligand (resiquimod or R848) [18], provided by the manufacturer, for validation purposes and as a positive control. Cells of the control and convalescent blood donors were stimulable to a similar extent when using LyoSpheres (Figure S3). As the assay was originally designed and optimized for the use of LyoSpheres and the binding of the containing ligands leads to a very strong cellular immune response, we detected a much higher IFN-*γ* release when compared to the stimulation with the specific SARS-CoV-2 peptides. However, as this had no direct impact on the significance of our results, there is no need to use LyoSpheres in similarly designed studies in the future.

While the immune cells of convalescent donors secreted increased amounts of IFN-*γ* after stimulation with SARS-CoV-2 specific peptides, the cells of control donors did not respond significantly to the specific SARS-CoV-2 peptide pool at all. This is in accordance with data published previously and, therefore, confirms the validity of the assay. Le Bert et al. showed an increased IFN-*γ* release after peptide stimulation in 36 individuals tested, who all recovered from COVID-19. In contrast to our study, the authors isolated peripheral blood mononuclear cells (PBMCs) and stimulated them with synthetic peptides that covered only the viral N protein. The measurement of IFN-*γ* release was performed with the ELISpot system mentioned above [19].

Our data suggest that cellular memory to SARS-CoV-2 is stably maintained for at least eight months after the COVID-19 disease and that the amount of IFN-*γ* release after peptide stimulation does not seem to correlate to the time post symptom onset. These results substantiate previous findings from other research groups that identified specific memory B [20] and T [17] cells eight and six months, respectively, after SARS-CoV-2 infection. We further observed no correlation between the cellular and humoral immune responses of convalescent donors. In detail, neither semiquantitative (Euroimmun, r = 0.2831) nor quantitative (DiaSorin, r = 0.2578) anti-SARS-CoV-2 IgG assay results correlated with the respective immune cell IFN-*γ* release after stimulation with SARS-CoV-2-specific peptides. All convalescents were seropositive at first donation; however, a successive antibody decay was observed for most donors. Previous studies had already suggested an early decay or even seroreversion of IgG antibodies in individuals with a mild disease progression [21, 22]. Nevertheless, IgG antibodies were still detectable in 95 % of all donors (39/41) included in our study on the day of the last donation and, therefore, up to eight months after symptom onset. No or only equivocal anti-SARS-CoV-2 IgA antibodies were detectable in some convalescents (8/41, 20 %) on the day of the last donation, whereby some were already seronegative at first donation (6/41, 15 %).This accords with the results of previous studies, which postulated a comparably fast decay of this antibody class after SARS-CoV-2 infection [10, 11]. However, an increased rate of false-positive results must be assumed due to the comparatively low specificity (71.5 % [*23*]) postulated for the Euroimmun anti-SARS-CoV-2 IgA assay used.

We further conducted a bead-based multiplex assay to determine more precisely against which viral antigens the anti-SARS-CoV-2 antibodies are targeted. We were able to elicit that all except one convalescent proband formed antibodies against each of the S domains examined (S1, S2 and RBD domain). This is important information because it suggests broad-spectrum antibody expression, which increases the potential for at least partial antibody binding to occur in spite of viral mutations. While nearly all convalescent donors formed antibodies against the S protein, only 62.5 % expressed antibodies against the viral N. Although data for the new coronavirus are lacking, different studies suggest that people infected with the SARS-CoV virus that appeared in 2004 were almost all positive for the expression of N protein-specific antibodies, whereas only about half of the convalescents expressed S protein-specific antibodies [24–26]. It is quite possible that the convalescents included in our study, who are apparently negative for N protein-specific antibodies, in fact, express antibodies which are not directed against the N-specific antigens coated on the microbeads of the assay used. Furthermore, the expression of antibodies against the viral N protein correlated well with the anti-SARS-CoV-2 IgG ratios determined in the serological Euroimmun assay (which detects antibodies against the viral S protein). This suggests a simultaneous degradation of N and S antibodies; whereby S antibodies may be detectable longer due to a higher initial titer.

As expected, no anti-SARS-CoV-2 antibodies were detected for most donors of the control cohort, however, antibodies against the viral S protein were identified for one control using the multiplex assay. This result initially suggested cross-reactivity, as no antibodies were detected for the residual targets. This was confirmed, as the immunological cells of this donor were not stimulable to release IFN-*γ* with the specific SARS-CoV-2 peptides. It is notable that the donor was also weakly positive in the remaining two anti-SARS-CoV-2 IgG serological assays, which are both designed for the detection of antibodies directed against the viral S protein. False-positive results may be due to the specificity of both serologic assays evaluated being less than 100 % [27]. It is interesting that the blood cells of the only convalescent donor for whom no antibodies against the S1, S2 and RBD domain were detectable in the multiplex assay were stimulable by SARS-CoV-2-specific peptides. For this donor, only equivocal seropositive IgG levels were detected in the two remaining seroassays. These examples suggest that especially SARS-CoV-2 infections which are longer past can be identified most reliably by measuring of IFN-y release after immune cell stimulation. To substantiate this, the main focus in follow-up studies should be on characterizing the cellular immunity of convalescents who initially expressed no or few antibodies or showed a rapid antibody decay.

Interestingly, neutralizing antibodies were detectable in all convalescent donors, whereas levels in all controls were below the cutoff, as expected. The values of the neutralizing antibodies correlated well with the quantitative (r = 0.7778) and semiquantitative (r = 0.7526) anti-SARS-CoV-2 IgG results obtained in the DiaSorin and Euroimmun assays, respectively. These results match those of Wajnberg et al., who detected neutralizing antibodies in a large proportion (> 90 %) of 30,082 individuals tested with mild to moderate disease progression. Neutralizing antibodies were thereby also persistent for months and neutralizing antibody levels also correlated well with the anti-SARS-CoV-2 IgG antibody results determined [12].

We next characterized immunological cell subpopulations of control and convalescent donors. The only difference we observed concerned the occurrence of NK cells, which was increased significantly in convalescents. This is a very interesting finding, as previous studies suggested that NK cells tend to be decreased in COVID-19 patients [28–30]. Most of these were patients with a severe course of COVID-19, whereas our study involves individuals with mild to moderate COVID-19 disease. Decreased NK cell levels in severe COVID-19 cases are associated with insufficient degradation of infected cells, which, *inter alia*, results in elevated inflammation [31]. Increased NK cell counts in individuals with mild progression could probably, therefore, contribute to avert a “cytokine storm,” that is known to trigger a severe COVID-19 outcome [32]. Characterization of leucocyte subpopulations in donor blood was performed without stimulation with anti-SARS-CoV-2-specific peptides. Because NK cells are IFN-*γ*-secreting [33] immunological memory cells [34, 35] the increased IFN-*γ* release of immune cells in the blood of convalescents after peptide stimulation could also be explained by an inductive formation of NK cells. However, this assumption needs further research on the involvement of NK cells in COVID-19 pathogenesis.

In summary, we successfully adapted a commercial assay to determine the cellular immune response in convalescent COVID-19 patients in a comparatively easy and fast way. Our data show that the newly developed assay identifies convalescent individuals reliably and is superior in its specificity to the serological assays used in this study. The cellular SARS-CoV-2-specific IFN-*γ* release assay described here might be helpful for the interpretation of the specificity of anti-SARS-CoV-2 antibodies detected especially when experienced COVID-19 disease is suspected but PCR-confirmed SARS-CoV-2 infection is missing. Our findings are of high importance because, compared to other studies, the cohort studied here includes convalescents with a mild disease course and a comparatively high mean age of 54 ± 8.4 years. Our results, therefore, also assume a long-lasting immunity against SARS-CoV-2 infection in the elderly.

## Data Availability

The datasets generated during the current study are available from the corresponding author on reasonable request.

## Abbreviations

SARS-CoV-2: Severe acute respiratory syndrome coronavirus type 2
COVID-19: Coronavirus Disease 2019
IFN-*γ*: Interferon-*γ*

## Literature

1. Khan S, Siddique R, Bai Q, Shabana Liu Y, Xue M, et al. Coronaviruses disease 2019 (COVID-19): Causative agent, mental health concerns, and potential management options. J Infect Public Health 2020;13(12):1840–4. doi:10.1016/j.jiph.2020.07.010

2. Cucinotta D, Vanelli M. WHO Declares COVID-19 a Pandemic. Acta Biomed. 2020;91(1):157–60. doi:10.23750/abm.v91i1.9397

3. Bönisch S, Wegscheider K, Krause L, Sehner S, Wiegel S, Zapf A, et al. Effects of coronavirus disease (COVID-19) related contact restrictions in Germany, March to May 2020, on the mobility and relation to infection patterns. Front Public Health 2020;8:568287. doi:10.3389/fpubh.2020.568287

4. Shereen MA, Khan S, Kazmi A, Bashir N, Siddique R. COVID-19 infection: Origin, transmission, and characteristics of human coronaviruses. J Adv Res. 2020;24:91–8. doi:10.1016/j.jare.2020.03.005

5. Koyama T, Platt D, Parida L. Variant analysis of SARS-CoV-2 genomes. Bull World Health Organ. 2020;98(7):495–504. doi:10.2471/BLT.20.253591

6. V’kovski P, Kratzel A, Steiner S, Stalder H, Thiel V. Coronavirus biology and replication: implications for SARS-CoV-2. Nat Rev Microbiol. 2020 Oct 28;1–16. doi:10.1038/s41579-020-00468-6

7. Salzberger B, Buder F, Lampl B, Ehrenstein B, Hitzenbichler F, Holzmann T, et al. Epidemiology of SARS-CoV-2 [in German]. Infection 2020 Aug;61(8):782–8. doi:10.1007/s15010-020-01531-3

8. Bošnjak B, Stein SC, Willenzon S, Cordes AK, Puppe W, Bernhardt G, et al. Low serum neutralizing anti-SARS-CoV-2 S antibody levels in mildly affected COVID-19 convalescent patients revealed by two different detection methods. Cell Mol Immunol. 2020 Nov 2;1–9. doi:10.1038/s41423-020-00573-9

9. Marklund E, Leach S, Axelsson H, Nyström K, Norder H, Bemark M, et al. Serum-IgG responses to SARS-CoV-2 after mild and severe COVID-19 infection and analysis of IgG non-responders. PLoS One 2020;15(10):e0241104. doi:10.1371/journal.pone.0241104

10. Isho B, Abe KT, Zuo M, Jamal AJ, Rathod B, Wang JH, et al. Persistence of serum and saliva antibody responses to SARS-CoV-2 spike antigens in COVID-19 patients. Sci Immunol. 52020 Oct 8;5(52):eabe5511. doi:10.1126/sciimmunol.abe5511

11. Iyer AS, Jones FK, Nodoushani A, Kelly M, Becker M, Slater D, et al. Persistence and decay of human antibody responses to the receptor binding domain of SARS-CoV-2 spike protein in COVID-19 patients. Sci Immunol. 2020 Oct 8;5(52):eabe0367. doi:10.1126/sciimmunol.abe0367

12. Wajnberg A, Amanat F, Firpo A, Altman DR, Bailey MJ, Mansour M, et al. Robust neutralizing antibodies to SARS-CoV-2 infection persist for months. Science 2020 Dec 4;370(6521):1227–30. doi:10.1126/science.abd7728

13. Thevarajan I, Nguyen THO, Koutsakos M, Druce J, Caly L, van de Sandt CE, et al. Breadth of concomitant immune responses prior to patient recovery: a case report of non-severe COVID-19. Nat Med. 2020;26(4):453–5. doi:10.1038/s41591-020-0819-2

14. Sekine T, Perez-Potti A, Rivera-Ballesteros O, Strålin K, Gorin J-B, Olsson A, et al. Robust T cell immunity in convalescent individuals with asymptomatic or mild COVID-19. Cell 2020;183(1):158-68.e14. doi:10.1016/j.cell.2020.08.017

15. Kuri-Cervantes L, Pampena MB, Meng W, Rosenfeld AM, Ittner CAG, Weisman AR, et al. Comprehensive mapping of immune perturbations associated with severe COVID-19. Sci Immunol. 2020 Jul 15;5(49):eabd7114. doi:10.1126/sciimmunol.abd7114

16. Laing AG, Lorenc A, Del Molino Del Barrio I, Das A, Fish M, Monin L, et al. A dynamic COVID-19 immune signature includes associations with poor prognosis. Nat Med. 2020 Oct;26(10):1623– 35. doi:10.1038/s41591-020-1038-6

17. Zuo J, Dowell A, Pearce H, Verma K, Long HM, Begum J, et al. Robust SARS-CoV-2-specific T-cell immunity is maintained at 6 months following primary infection. 2020. doi:10.21203/rs.3.rs-101480/v1

18. Yong MK, Cameron PU, Spelman T, Elliott JH, Fairley CK, Boyle J, et al. Quantifying adaptive and innate immune responses in HIV-infected participants using a novel high throughput assay. PLoS One 2016 Dec 9;11(12):e0166549. doi:10.1371/journal.pone.0166549

19. Le Bert N, Tan AT, Kunasegaran K, Tham CYL, Hafezi M, Chia A, et al. SARS-CoV-2-specific T cell immunity in cases of COVID-19 and SARS, and uninfected controls. Nature 2020;584(7821):457– 62. doi:10.1038/s41586-020-2550-z

20. Hartley GE, Edwards ESJ, Aui PM, Varese N, Stojanovic S, McMahon J, et al. Rapid generation of durable B cell memory to SARS-CoV-2 spike and nucleocapsid proteins in COVID-19 and convalescence. Sci Immunol. 2020;5(54):eabf8891. doi:10.1126/sciimmunol.abf8891

21. Ibarrondo FJ, Fulcher JA, Goodman-Meza D, Elliott J, Hofmann C, Hausner MA, et al. Rapid decay of anti-SARS-CoV-2 antibodies in persons with mild Covid-19. N Engl J Med. 2020;383(11):1085– 7. doi:10.1056/NEJMc2025179

22. Röltgen K, Wirz OF, Stevens BA, Powell AE, Hogan CA, Najeeb J, et al. SARS-CoV-2 antibody responses correlate with resolution of RNAemia but are short-lived in patients with mild illness. medRxiv 2020 Aug 17. doi:10.1101/2020.08.15.20175794

23. Manalac J, Yee J, Calayag K, Nguyen L, Patel PM, Zhou D, et al. Evaluation of Abbott anti-SARS-CoV-2 CMIA IgG and Euroimmun ELISA IgG/IgA assays in a clinical lab. Clin Chim Acta 2020;510:687–90. doi:10.1016/j.cca.2020.09.002

24. Huang L-R, Chiu C-M, Yeh S-H, Huang W-H, Hsueh P-R, Yang W-Z, et al. Evaluation of antibody responses against SARS coronaviral nucleocapsid or spike proteins by immunoblotting or ELISA. J Med Virol. 2004;73(3):338–46. doi:10.1002/jmv.20096

25. Wang Y, Chang Z, Ouyang J, Wei H, Yang R, Chao Y, et al. Profiles of IgG antibodies to nucleocapsid and spike proteins of the SARS-associated coronavirus in SARS patients. DNA Cell Biol. 2005;24(8):521–7. doi:10.1089/dna.2005.24.521

26. Leung DTM, Tam FCH, Ma CH, Chan PKS, Cheung JLK, Niu H, et al. Antibody response of patients with severe acute respiratory syndrome (SARS) targets the viral nucleocapsid. J Infect Dis. 2004;190(2):379–86. doi:10.1086/422040

27. Manthei DM, Whalen JF, Schroeder LF, Sinay AM, Li S-H, Valdez R, et al. Differences in performance characteristics among four high-throughput assays for the detection of antibodies against SARS-CoV-2 using a common set of patient samples. Am J Clin Pathol. 2020 Oct 9;aqaa200. doi:10.1093/ajcp/aqaa200

28. Zheng M, Gao Y, Wang G, Song G, Liu S, Sun D, et al. Functional exhaustion of antiviral lymphocytes in COVID-19 patients. Cell Mol Immunol. 2020;17(5):533–5. doi:10.1016/j.cell.2018.11.048

29. Li D, Chen Y, Liu H, Jia Y, Li F, Wang W, et al. Immune dysfunction leads to mortality and organ injury in patients with COVID-19 in China: insights from ERS-COVID-19 study. Signal Transduct Target Ther. 2020 May 5;5(1):62. doi:10.1038/s41392-020-0163-5

30. Wang F, Nie J, Wang H, Zhao Q, Xiong Y, Deng L, et al. Characteristics of peripheral lymphocyte subset alteration in COVID-19 pneumonia. J Infect Dis. 2020;221(11):1762–9. doi:10.1093/infdis/jiaa150

31. van Eeden C, Khan L, Osman MS, Cohen Tervaert JW. Natural killer cell dysfunction and its role in COVID-19. Int J Mol Sci. 2020 Sep 1;21(17):6351. doi:10.3390/ijms21176351

32. Coperchini F, Chiovato L, Croce L, Magri F, Rotondi M. The cytokine storm in COVID-19: An overview of the involvement of the chemokine/chemokine-receptor system. Cytokine Growth Factor Rev. 2020;53:25–32. doi:10.1016/j.cytogfr.2020.05.003

33. Paolini R, Bernardini G, Molfetta R, Santoni A. NK cells and interferons. Cytokine Growth Factor Rev. 2015;26(2):113–20. doi:10.1016/j.cytogfr.2014.11.003

34. Sun JC, Lanier LL. Is there natural killer cell memory and can it be harnessed by vaccination? NK cell memory and immunization strategies against infectious diseases and cancer. Cold Spring Harb Perspect Biol. 2018 Oct1;10(10):a029538. doi:10.1101/cshperspect.a029538

35. Brillantes M, Beaulieu AM. Memory and memory-like NK cell responses to microbial pathogens. Front Cell Infect Microbiol. 2020;10:102. doi:10.3389/fcimb.2020.00102

