## Supplement for "Evidence of long-lasting humoral and cellular immunity against SARS-CoV-2 even in elderly COVID-19 convalescents showing a mild to moderate disease progression"

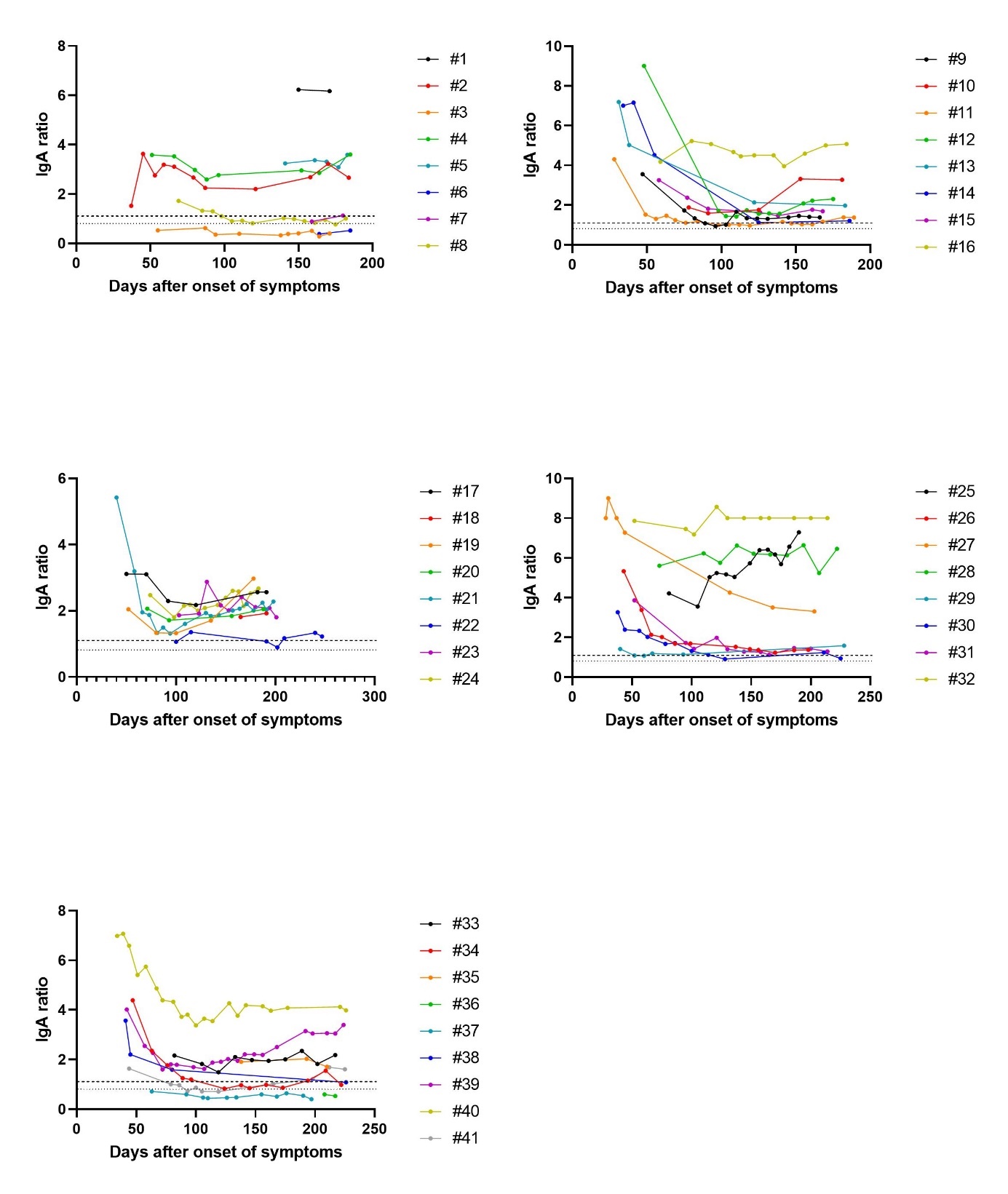

**Figure S1: Persistence and decay of anti-SARS-CoV-2 IgA antibodies in COVID-19 convalescents (n = 41).** Anti-SARS-CoV-2 IgA antibody expression of convalescents on the days after onset of symptoms indicated. Semiquantitative ratios were determined using the ELISA-based Euroimmun assay. According to the manufacturer, values above the upper dashed line (ratio = 1.1) are considered seropositive and values below the lower dashed line (ratio = 0.8) are considered seronegative. Values between the horizontal dashed lines were interpreted as equivocal-positive.

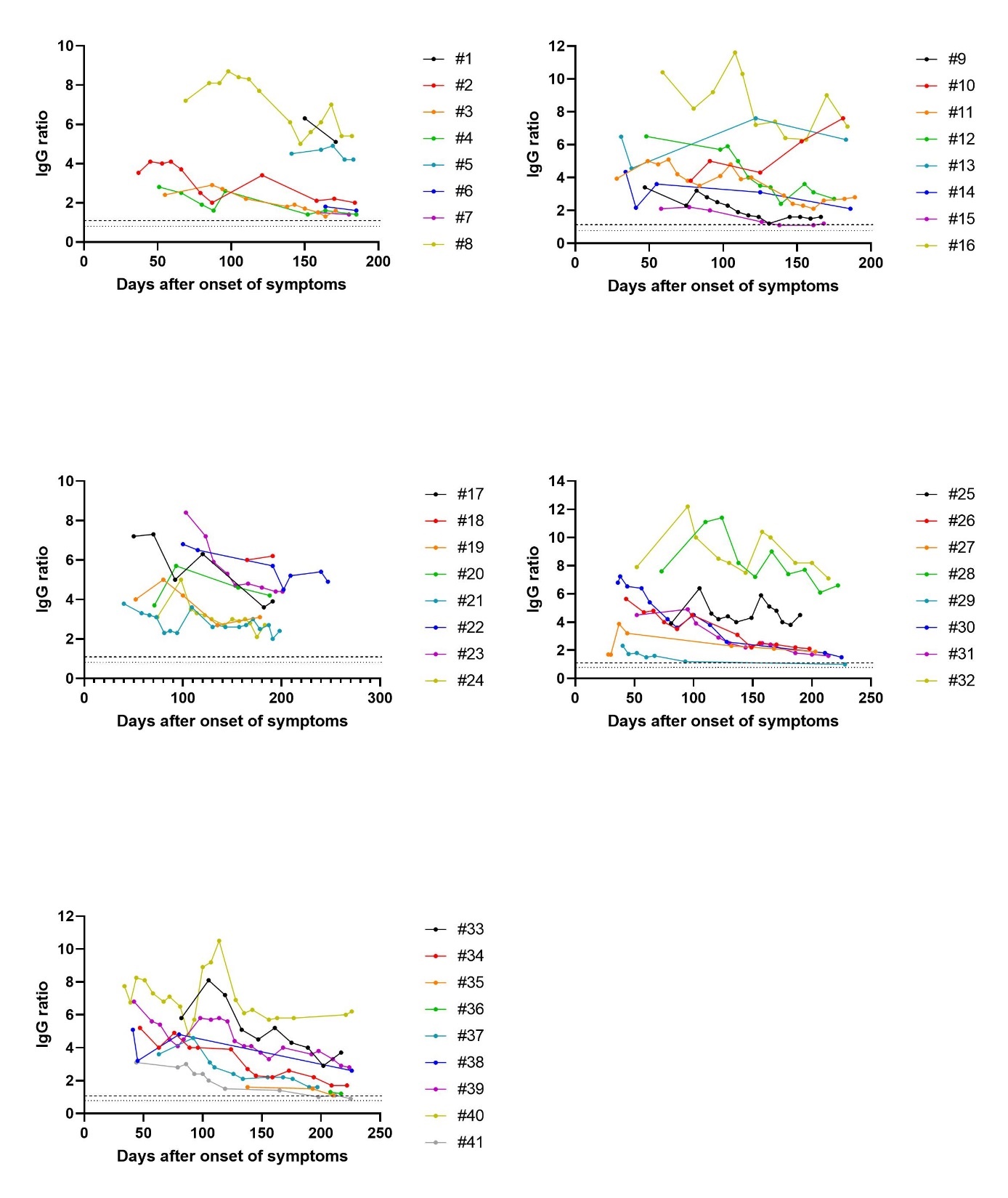

**Figure S2: Persistence and decay of anti-SARS-CoV-2 IgG antibodies in COVID-19 convalescents (n = 41).** Anti-SARS-CoV-2 IgG antibody expression of convalescents on the days after onset of symptoms indicated. Semiquantitative ratios were determined using the ELISA-based Euroimmun assay. According to the manufacturer, values above the upper dashed line (ratio = 1.1) are considered seropositive and values below the lower dashed line (ratio = 0.8) are considered seronegative. Values between the horizontal dashed lines were interpreted as equivocal-positive.

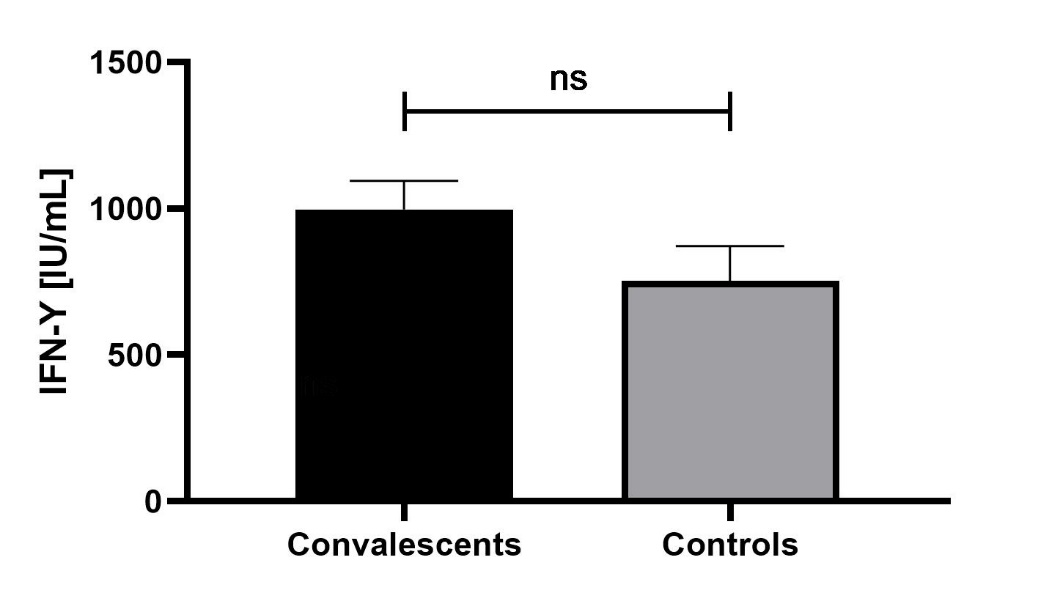

**Figure S3: IFN-γ release after stimulation of whole blood of convalescents (n = 41) and controls (n = 18) with LyoSpheres pellets.** Cells in our study were stimulated in a parallel setup using LyoSpheres pellets containing a CD3 T-cell receptor agonist and a viral TLR 7/8 ligand (resiquimod or R848) for assay-validation and as a positive control. A comparatively high IFN-γ release was detected because the QuantiFeron-assay was originally designed and optimized for the use of LyoSpheres. Nevertheless, no significant differences in the IFN-γ secretion was detected after stimulation of the whole blood of COVID-19 convalescents and healthy controls. ns: not significant.
